# Symptom-based ordinal scale fuzzy clustering of functional gastrointestinal disorders

**DOI:** 10.1101/2020.05.11.20098376

**Authors:** Marjan Mansoruian, Hamid Reza Marateb, Ammar Hassanzadeh Keshteli, Hamed Daghagh Zadeh, Miquel Angel Mañanas, Peyman Adibi

**Author notes:** Address for correspondence: Hamid Reza Marateb Biomedical Engineering Research Center (CREB), Building H, Floor 4, Av. Diagonal 647, 08028 Barcelona, Spain, https://bioart.upc.edu/en/staff/.

## Abstract

**Background:** The validity of Rome III criteria for diagnosing functional gastrointestinal disorders (FGIDs) have been frequently questioned in the literature. In epidemiology, when a disease is diagnosed, the existence of a true cluster must be proven. Thus, clustering the common GI symptoms of individuals and comparing the clusters with FGIDs defined by the Rome III criteria could provide insights about the validity of FGIDs defined by those criteria. Well-separated compact clusters were detected in responses to questionnaires of the epidemiological features of different FGIDs in Iranian adults using fuzzy ordinal clustering. The representative sample from each cluster i.e. Cluster Representative (CR) was formed whose corresponding FGID was diagnosed with Rome III criteria. Then, FGID diagnosis was performed for all participants in each cluster and the percentage of cases whose FGID was the same as the cluster’s identified FGID (agreement) was reported.

**Results:** Fourteen valid clusters were detected in 4763 people. The average membership of the objects in each cluster was 77.3%, indicating similarity of the objects in clusters to their corresponding CRs. Eight clusters were assigned to single FGIDs (irritable bowel syndromes: constipation IBS-C, diarrhea IBS-D and un-subtyped IBS-U; functional bloating FB; functional constipation FC; belching disorder BD. The agreement was higher than 50% in single FGID clusters except those whose diagnosis was IBS-U.

**Conclusions:** IBS-C, IBS-D, FC, BD, and FB defined with Rome III criteria exist in the population, which is not the case for IBS-U.

## Introduction

Throughout documented history, structural diseases could be identified by pathologists and at times cured by medical technology. However, the nonstructural symptoms described as “functional” remain enigmatic and less amenable for explanation or effective treatment (1).

The research consistency and more reliable diagnosis of FGIDs have been attained utilizing the Rome criteria in conjunction with self-reported questionnaires (1, 2). The Rome classification system is based on symptom clusters assumed to remain consistent through population groups. These criteria have been modified occasionally, and have undergone many revisions since their first presentation (3).

A number of studies have been performed in the literature on the validity of symptom-based FGID diagnosis(4-7). Eslick *et al* performed the factor analysis of 46 symptom items of 897 patients to identify the underlying latent factors. A *k*-means cluster analysis was then applied on the extracted factors to detect the number of patient groups (4). Alessandro Digesu *et al* examined whether Rome III criteria are reliable to diagnose constipation in 201 women. They found out that the Rome III questionnaire did not have significant construct validity with patients’ self-report of constipation, stool frequency and stool form. They finally concluded that Rome III is not a valid instrument to diagnose constipation(6). Whitehead and Drossman performed a critical review on the validity of symptom-based criteria for diagnosing irritable bowel syndrome (IBS). They concluded that the sensitivity and specificity of those criteria are 40%-90% and 70%-90%, respectively with clinical diagnoses made by experienced clinicians (7). Dang *et al* performed a systematic review of symptom-based diagnostic criteria for IBS. They considered different validation studies. The patient sample size was less than 350 in any studies considered. Based on the manuscripts reviewed by the authors until 2019, they demonstrated poor validity and utilization of Rome III for diagnosing IBS(5).

Symptom-based diagnosis of distinct FGIDs has been criticized on epidemiology due to the overlap between symptoms(4). Traditional clustering methods such as *k*-means or hierarchical algorithms do not allow the overlap between symptoms or FGIDs. This problem has been solved in data mining by using fuzzy clustering(8). Using advanced Fuzzy data-mining techniques on large sample sizes, could give new insights on the validity of symptom-based FGID diagnosis.

This study aimed to identify whether FGIDs could be identified based on common gastrointestinal symptoms of individuals who formed compact and well-separated epidemiological groups (clusters) in a population.

## Methods

The Study on the Epidemiology of Psychological, Alimentary Health and Nutrition (SEPAHAN) project was designed to be a two-phase cross-sectional study. All information was gathered through self administered questionnaires mainly aimed to investigate the epidemiological features of different FGIDs within a large sample of Iranian adults working in 50 different healthcare centers across Isfahan province, Iran (9). The 4-item rating ordinal-measurement scale including “never or rarely”,” sometimes”,” often”, and “always” labeled as 1-4, were used in our analysis.

Overall, detailed information from 4763 individuals became available for further analysis. The subjects did not have any alarm symptoms. All subjects gave informed consent to the experimental procedure. Ethical approval to conduct the study was provided by the Medical Research Ethics Committee of IUMS (#189069, #189082, and #189086), conformed to the Declaration of Helsinki.

The main research question in our study was as follow: Do FGIDs diagnosed using Rome III criteria have fuzzy ontology? Also, two research hypotheses were taken into account in our study. 1) It is possible to accurately determine subject groups based on their responses to GI questionnaire; and 2) The groups formed by the clustering method could be assigned to one/more FGIDs, based on the Rome III criteria. Thus, an unsupervised machine learning approach was developed for the accurate identification of different groups (clusters) of FGIDs according to SEPAHAN questionnaire items. The particle swarm optimization-based unsupervised-fuzzy clustering technique was used as the optimum ordinal-to-interval measurement scale conversion.

Data were analyzed using Matlab and Statistics Toolbox Release 2012a (The MathWorks, Inc., Natick, Massachusetts, USA). Quantitative data were expressed as mean (SD) and qualitative as number (percent). Determining different FGID clusters according to SEPAHAN questionnaire using the clustering analysis, 31 significant head questions were selected by two experts in gastroenterology.

First, those whose response to *all* of the 31 selected head questions was either “never or rarely” or “ sometimes” were regarded as “healthy” and excluded from the clustering analysis. Thus, the following clustering methodology is related to unhealthy subjects. The matrix (“IN”) was then formed for the unhealthy subjects, in which the labels of the answered questions for each person were regarded as a row.

People having similar symptoms could belong to distinct diagnostic groups (FGID categories). Fuzzy clustering could be enrolled in which each subject belongs to all of the FGID groups (clusters) with different membership degrees in [0, 1]. This gives the flexibility to express that objects can belong to more than one cluster. In fact, the terms ‘cluster’ and ‘cluster membership’ are identical with the epidemiological terms ‘group’ and ‘group membership’ (4). Also, when a disease is diagnosed in epidemiology, the existence of a true disease cluster must be proven(10). The clustering algorithm is briefly illustrated as **Algorithm 1** in the appendices. The clustering method used in our study has been presented and then validated on ordinal datasets in the literature and showed promising performance by Brouwer and Groenwold (11) and Marateb *et al* (12).

A good partition produces a small value for the compactness, while well-separated clusters produce a high value for the separation. Thus, the best partition is related to the minimum of the XB CVI defined as the ratio of compactness to separation (13). After detecting the best clustering structure, similar clusters were merged using single-linkage clustering (using the cluster representatives as the input samples)(14). Each subject was then classified to the cluster for which its membership value was the highest. Like many epidemiologic studies in which a sample object is required from a target population of interest(15), we defined the cluster representative of each group as the representative sample (object) from that cluster. The representative sample was chosen from the objects of corresponding cluster, where the sum of the distance of the other objects in that cluster to the representative is minimal. The representative sample was, in fact, the vector of representative symptoms. The symptoms were then compared with those of ROME III by an expert in Gastroenterology and the corresponding FGIDs were identified.

## Results

The socio-demographic and anthropometric factors of the sample population are listed in Table 1. The percentage of healthy people, as defined in the statistical methods section, was 51.15%. They were excluded from the descriptive and clustering analysis.

**Table 1:** General characteristics of study participants across different categories of variables

| Variables | categories | Descriptive statistics |
| --- | --- | --- |
| **Sex** | Male | 2106(%44.2) |
|  | Female | 2657(%55.8) |
| **Marital status** | Single | 3776(%81.2) |
|  | Married | 795(%17.1) |
|  | Divorced or widowed | 79 (1.7%) |
| **Educational level** | under diploma | 638 (13.4%) |
|  | Diploma | 1348 (28.3%) |
|  | above diploma but under master | 2338 (49.1%) |
|  | master or above | 312 (6.6%) |
| **Age (year)** | - | 36.6±3.1 |
| **BMI (kg/m<sup>2</sup>)** | - | 24.9±5.1 |
| **Weight (kg)** | - | 68.89±13.49 |

All values are mean ± SD for quantitative variables and number (percent for qualitative variables; BMI: Body mass index;

The average membership of the objects in each cluster is shown in Table 2. The membership value for each object was calculated in terms of the relative similarity (16) between the 31 ordinal-scale questionnaire answers of that subject and those of the corresponding cluster representative. It ranged from 0% (complete dissimilarity) to 100% (complete similarity or clustering consistency).

**Table 2:** The identified functional gastrointestinal disorders subtypes and the percentage of people in each cluster, and the average membership of the objects in each cluster identified in the SEPAHAN dataset shown in mean ± SD.

| Cluster Index | C1 | C2 | C3 | C4 | C5 | C6 | C7 | C8 | C9 | C10 | C11 | C12 | C13 | C14 |
| --- | --- | --- | --- | --- | --- | --- | --- | --- | --- | --- | --- | --- | --- | --- |
| % People <sup>a</sup> | 1.46 | 2.58 | 1.41 | 0.61 | 2.48 | 0.98 | 6.12 | 8.80 | 2.08 | 1.14 | 8.45 | 8.11 | 4.41 | 0.22 |
| Average membership <sup>b</sup> (%) | 83±5 | 82±6 | 72±6 | 56±8 | 79±5 | 77±5 | 83±8 | 82±10 | 77±5 | 73±6 | 82±10 | 83±7 | 78±8 | 75±5 |
| FGIDs <sup>c</sup> | IBS-C<br>+ FD<br>(non<br>PDS) | IBS-<br>U | IBS-C<br>+ FD<br>(PDS) | IBS-C+<br>FD<br>(PDS)+<br>UH | IBS-<br>C+ FD<br>(PDS) | IBS-<br>D | FB | FB | IBS-<br>C | IBS-C+<br>FD<br>(PDS) | BD | IBS-<br>U | FC | IBS-<br>M+<br>UH |

<sup>a</sup>: The total percentage of people is 48.9%. They are the “unhealthy” people in the analyzed samples. The rest, i.e. “healthy” people were excluded from the analysis as illustrated in the “statistical methods” section. The percentage of people was calculated as calculated as the number of people in each cluster divided by the total sample size of 4763. <sup>b</sup>: The membership value for each object was calculated in terms of the relative similarity between 31 the questionnaire answers of that subject and those of the corresponding cluster representative. <sup>c</sup>: Identification of common FGIDs was done by two gastroenterologists. FGIDs: functional gastrointestinal disorders, IBS: irritable bowel syndrome, IBS-C: constipation predominant IBS, IBS-D: diarrhea predominant IBS, IBS-M: mixed IBS, IBS-U: unsubtyped IBS, FD: functional dyspepsia, FB: functional bloating, FC: functional constipation, PDS: postprandial distress syndrome, UH: uninvestigated heartburn, BD: belching disorder.

The percentage of people (i.e., the number of people divided by 4763) belonging to identified clusters is also listed in Table 2. The matched FGIDs identified using the major symptoms of the subjects in the identified clusters are also shown in Table 2.

Clusters C2, C6, C7, C8, C9, C11, C12, and C13 were assigned to specific FGIDs based on the similarity of the cluster representative with Rome III criteria. The agreement between their symptoms and those of assigned FGIDs were shown as Venn diagrams in Figure 1. In this figure, symptom-based FGID diagnosis was performed for all participants in each cluster based on the Rome III criteria, and the percentage of cases whose FGID was the same as the cluster’s identified FGID was shown. The agreement was higher than the cut-off similarity level of 50% (17) in all cases except those whose diagnosis was IBS-U (C2, and C12). C2 and C12 were both assigned to IBS-U, although C12 had minor variations with C2. For example, stoppage, enema and defecation problems existed in C12 but not C2. In C2 cluster, people often had bloating which was not the case in C12. C7 and C8 were both assigned to functional bloating. Early satiety, borborygmi and bloating were more frequent in C8 while dyspepsia, belching, and abdominal/non-cardiac chest pain and fullness were more frequent in C7. There were insufficient criteria for the diagnosis of functional dyspepsia, IBS or other FGIDs. They could be considered as functional abdominal bloating based on Rome I and II criteria, although the name was changed to functional bloating in Rome III criteria (18).

**Figure 1:**
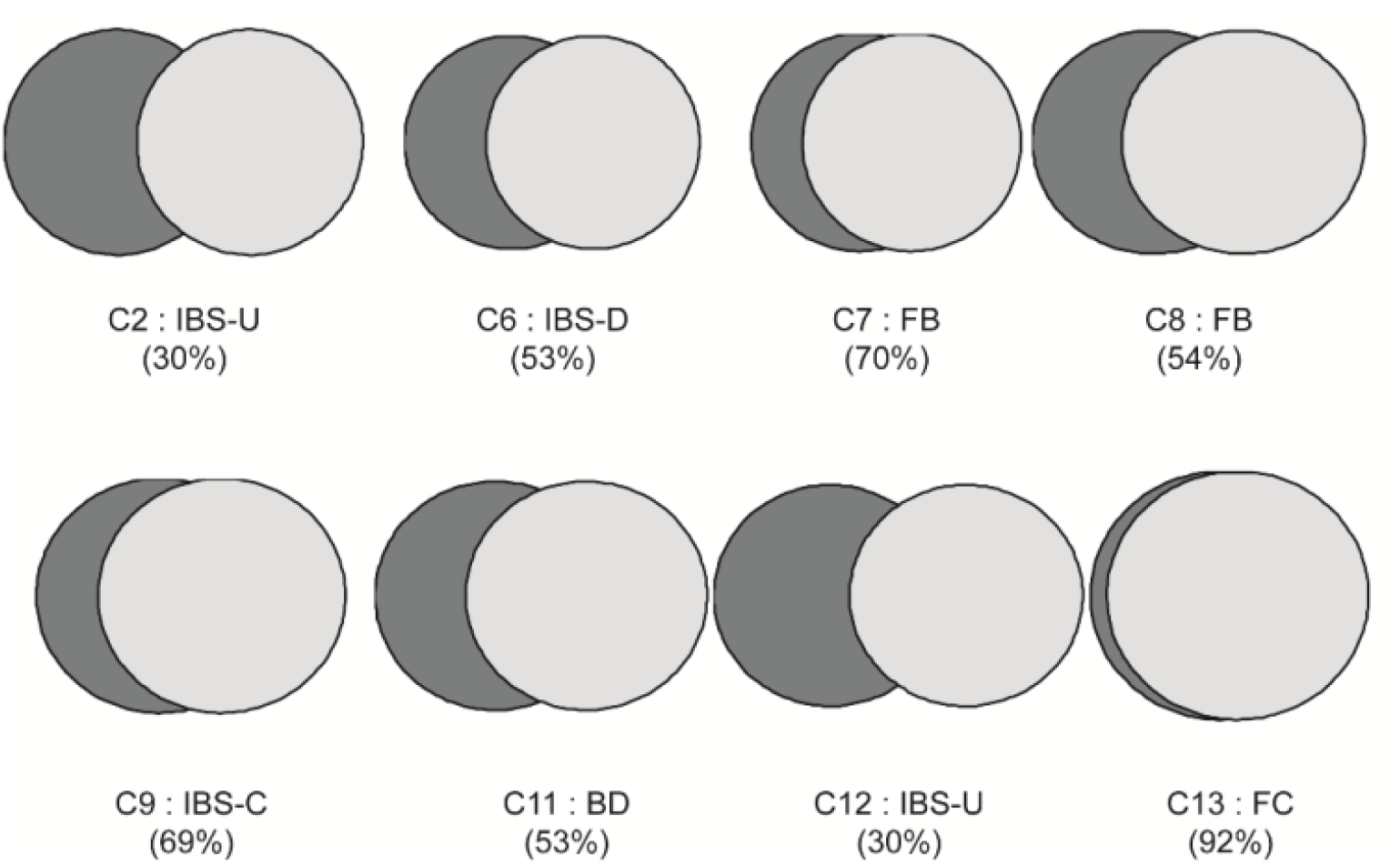
The overlap between identified clusters and corresponding functional gastrointestinal disorders (FGIDs) (Based on the Rome III criteria) shown as a Venn diagram. The percentage of the cases whose symptom-based diagnosis was identical to those of their cluster representatives were reported for clusters assigned to a single FGID. IBS: irritable bowel syndrome, IBS-C: constipation predominant IBS, IBS-D: diarrhea predominant IBS, IBS-U: unsubtyped IBS, FB: functional bloating, FC: functional constipation, PDS: postprandial distress syndrome, UH: uninvestigated heartburn, BD: belching disorder.

Cluster C3 was the combination of IBS-C (52%) and PDS (96%), while clusters C14 included IBS-M (43%) and uninvestigated heartburn (85%), where the agreement percentage was shown in the parenthesis. Cluster C1 was the combination of IBS-C (7%) and FD (non-PDS) (34%)). On the other hand, cluster C5 was the combination of IBS-C (1%) and FD (PDS) (74%). Clusters C4 and C10 had the combination IBS-C (24%, 38%), and functional dyspepsia (PDS) (100%, 85%). However, the symptoms of C10 were not as severe as those of C4. For example, anal sore, bloating, mucus, defecation problem, enema, incomplete evacuation, hardness, and abdominal pain were more frequent in C4 in comparison with C10. In the meanwhile, uninvestigated heartburn (21%) was also present in C4. Two identified clusters C4 and C10, are examples of the concept of severity in IBS introduced by Drossman *et al*(19).

## Discussion

The clustering algorithms have been widely used in clinical studies to identify subtypes of diseases or human disease etiological factors; this is a well-known methodology(20). In epidemiology, when a disease is first identified, its occurrence as a true cluster (group) must be proven(10). The validity of the identified clusters was assessed based on 1) XB CVI that ensures the clusters are compact and well-separated, and 2) the average membership (similarity) of the objects in each cluster (Table 2; above 77% in single FGID clusters). The cluster representative was selected similar to the epidemiological studies as the sample representative of that cluster(15). The existence of the FGIDs with the minimum similarity level of 50%(17), could be proven in our study. However, since the overlap (similarity) of the single-FGID clusters with Rome-III defined FGIDs are between 0 %-100 %, their degree of existence could be stated in the framework of Fuzzy ontology(21). Thus, the main research question was answered. As a validation of this hypothesis, in total, 21.07% of the population had IBS (Table 2). A recent patient-based analysis of the same database showed that the prevalence of IBS was 21.7% using Rome III diagnostic criteria (22).

Our results concerning that IBS-C, and D could be symptom-based diagnosed are in agreement with those of Whitehead *et al* (23) and Whitehead and Drossman (24). However, this is in contrast with some studies that claimed poor validity and utilization of Rome III criteria (4, 5, 25). Since the clustering was performed on all of the 31 significant head questions, high agreement with FGIDs diagnosed by few items in Rome III is very hard to reach. Thus, like many microbiological studies(17), the cut-off similarity value of 50% was used in our study. However, tightening the cut-off similarity level to about 70%, which is suitable for distinguishing unambiguous epidemiological disease, the ontology of the following FGIDs FC, FB and IBS-C could be verified(17).

In our study, each subject was finally assigned to a cluster. This is one of the limitations of our study in which concurrent FGIDs could not be entirely identified. Alternatively, the membership function could be used to identify different clusters to which a subject belongs that will be focused in the future.

## Data Availability

The datasets analyzed during the current study are not publicly available due to the institution limitations.

## Abbreviations

BD: belching disorder
CR: Cluster Representative
FGIDs: functional gastrointestinal disorders
FB: functional bloating
FC: functional constipation
IBS: irritable bowel syndromes
IBS: irritable bowel syndrome
IBS-C: constipation predominant IBS
IBS-D: diarrhea predominant IBS
IBS-U: unsubtyped IBS
PDS: postprandial distress syndrome
SEPAHAN: Study on the Epidemiology of Psychological, Alimentary Health and Nutrition
UH: uninvestigated heartburn

## Ethics Approval

Written informed consents were obtained from all participants, and the ethics committee of Isfahan University of medical sciences approved the study.

## Authors Contribution

M.M., HR.M., P.A, and A.H.K designed research; M.M. and HR.M. analyzed data; M.A.M and M.M. wrote the paper. All authors read and approved the final manuscript.

## Acknowledgements

The authors are grateful to Kevin McGill for reviewing a draft of this paper. The authors are thankful to Sobhan Goudarzi for collaboration in implementing part of the clustering algorithm.

## Appendices

### Algorithm 1: Identifying the clustering structure of the input matrix data “IN”

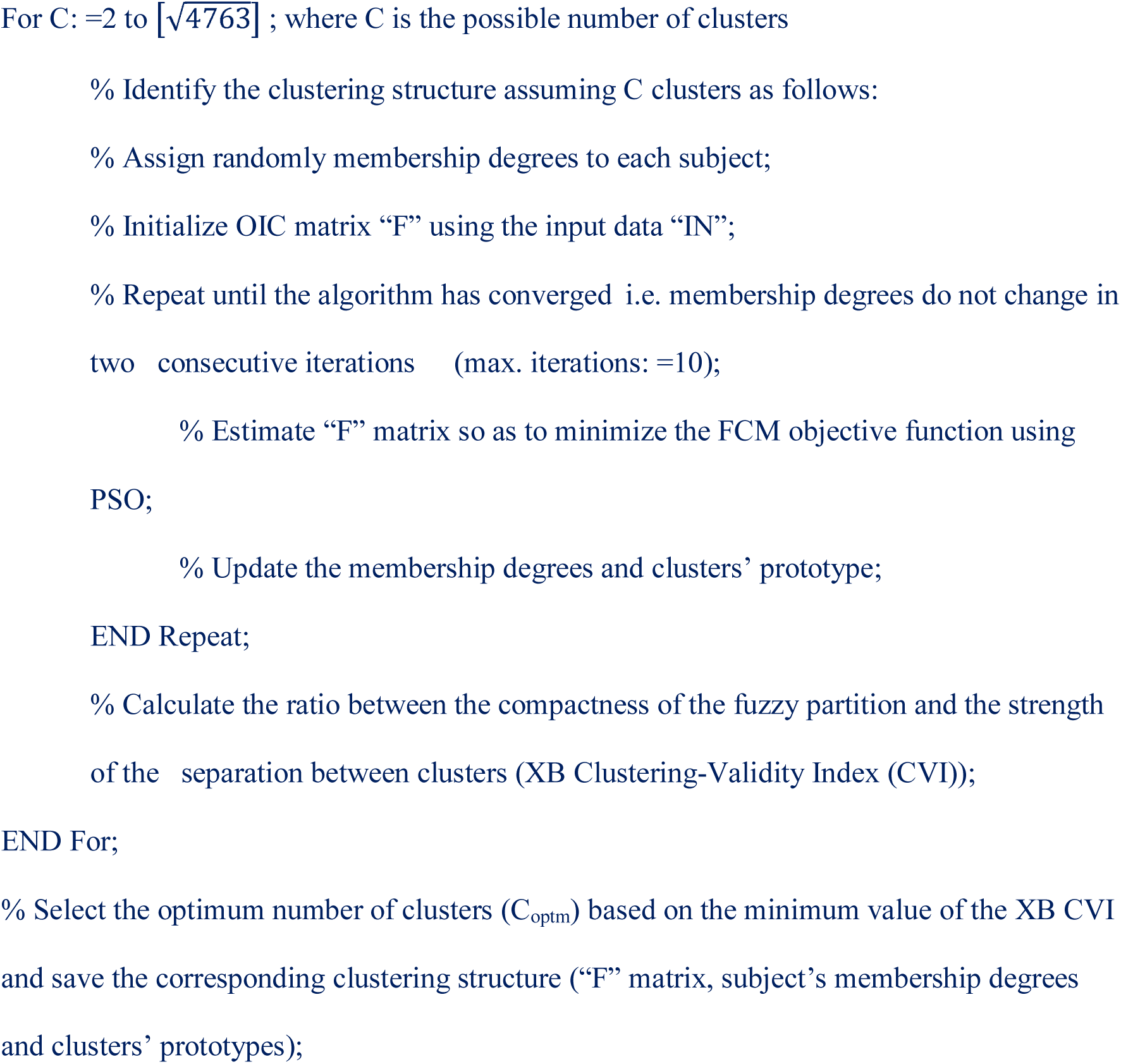

## References

1. Drossman DA. The functional gastrointestinal disorders and the Rome III process. Gastroenterology. 2006;130(5):1377–90.

2. Drossman DA, Dumitrascu DL. Rome III: New standard for functional gastrointestinal disorders. Journal of Gastrointestinal and Liver Diseases. 2006;15(3):237.

3. Jung HK. Rome III Criteria for Functional Gastrointestinal Disorders: Is There a Need for a Better Definition? Journal of neurogastroenterology and motility. 2011;17(3):211–2.

4. Eslick GD, Howell SC, Hammer J, Talley NJ. Empirically derived symptom sub-groups correspond poorly with diagnostic criteria for functional dyspepsia and irritable bowel syndrome. A factor and cluster analysis of a patient sample. Alimentary pharmacology & therapeutics. 2004; 19(1):133–40.

5. Dang J, Ardila-Hani A, Amichai MM, Chua K, Pimentel M. Systematic review of diagnostic criteria for IBS demonstrates poor validity and utilization of Rome III. Neurogastroenterology and motility: the official journal of the European Gastrointestinal Motility Society. 2012;24(9):853-e397.

6. Digesu GA, Panayi D, Kundi N, Tekkis P, Fernando R, Khullar V. Validity of the Rome III Criteria in assessing constipation in women. International urogynecology journal. 2010;21(10):1185–93.

7. Whitehead WE, Drossman DA. Validation of symptom-based diagnostic criteria for irritable bowel syndrome: a critical review. The American journal of gastroenterology. 2010;105(4):814–20; quiz 3, 21.

8. Szczepaniak PS, Lisboa PJ, Kacprzyk J. Fuzzy systems in medicine: Springer; 2000.

9. Adibi P, Keshteli AH, Esmaillzadeh A, Afshar H, Roohafza H, Bagherian-Sararoudi H, et al. The study on the epidemiology of psychological, alimentary health and nutrition (SEPAHAN): overview of methodology. J Res Med Sci. 2012;17(5).

10. Greenwald P, Block G. Methodological Issues in Epidemiological Studies of Disease Clusters and Environmental Contamination. In: Vouk VB, Butler GC, Upton AC, Parke DV, Asher SC, editors. Methods for assessing the effects of mixtures of chemicals. Stanford University: Wiley Chichester; 1987.

11. Brouwer RK, Groenwold A. Modified fuzzy c-means for ordinal valued attributes with particle swarm for optimization. Fuzzy Sets and Systems. 2010;161(13):1774–89.

12. Marateb HR, Mansourian M, Adibi P, Farina D. Manipulating measurement scales in medical statistical analysis and data mining: A review of methodologies. Journal of Research in Medical Sciences. 2014;19(1).

13. Kim D-W, Lee KH, Lee D. On cluster validity index for estimation of the optimal number of fuzzy clusters. Pattern Recognition. 2004;37(10):2009–25.

14. Duda RO, Hart PE, Stork DG. Pattern classification: John Wiley & Sons; 2012.

15. Rothman KJ, Greenland S, Lash TL. Modern epidemiology: Lippincott Williams & Wilkins; 2008.

16. Ichino M, Yaguchi H. Generalized Minkowski metrics for mixed feature-type data analysis. Systems, Man and Cybernetics, IEEE Transactions on. 1994;24(4):698–708.

17. Dijkshoorn L, Towner KJ, Struelens M. New approaches for the generation and analysis of microbial typing data. Amsterdam; New York: Elsevier; 2001. xi, 357 p. p.

18. Seo AY, Kim N, Oh DH. Abdominal Bloating: Pathophysiology and Treatment. Journal of neurogastroenterology and motility. 2013;19(4):433–53.

19. Drossman DA, Chang L, Bellamy N, Gallo-Torres HE, Lembo A, Mearin F, et al. Severity in irritable bowel syndrome: a Rome Foundation Working Team report. The American journal of gastroenterology. 2011;106(10):1749–59; quiz 60.

20. Armstrong RA, Wood L. The identification of pathological subtypes of Alzheimer’s disease using cluster analysis. Acta neuropathologica. 1994;88(1):60–6.

21. Sadegh-Zadeh K. Handbook of analytic philosophy of medicine. Dordrecht; New York: Springer Verlag; 2012. xxiv, 1133 p. p.

22. Esmaillzadeh A, Keshteli AH, Hajishafiee M, Feizi A, Feinle-Bisset C, Adibi P. Consumption of spicy foods and the prevalence of irritable bowel syndrome. World journal of gastroenterology: WJG. 2013;19(38):6465–71.

23. Whitehead W, Palsson O, Thiwan S, Talley N, Chey W, Irvine E, et al. Development and validation of the Rome III diagnostic questionnaire. Rome III: The Functional Gastrointestinal Disorders 3rd Edition ed McLean, VA: Degnon Associates, Inc. 2006: 835–53.

24. Whitehead WE, Drossman DA. Validation of Symptom-Based Diagnostic Criteria for Irritable Bowel Syndrome: A Critical Review. Am J Gastroenterol. 2010;105(4):814–20.

25. Digesu GA, Panayi D, Kundi N, Tekkis P, Fernando R, Khullar V. Validity of the Rome III Criteria in assessing constipation in women. International urogynecology journal. 2010;21(10):1185–93.

